# Health literacy, vaccine literacy, and lifestyle behaviours among young adults in Pakistan: A cross-sectional study

**DOI:** 10.64898/2026.06.30.26356971

**Authors:** Subboh Mushtaq, Kristina Motiejūnaitė

## Abstract

Health literacy (HL) is a critical determinant of health behaviours and outcomes. Vaccine literacy (VL), a domain-specific extension of HL, has emerged as an important determinant of vaccination attitudes. However, evidence examining the combined relationship of HL and VL with lifestyle behaviours remains limited, particularly among young adults in Pakistan. A cross-sectional study was conducted among 587 participants aged 18 years and above in Pakistan. HL was assessed using the European Health Literacy Population Survey 2019–2021 questionnaire (HLS19-Q; 17 items; 5-point Likert scale; score range 0–100) and VL was assessed using the HLS19 vaccine literacy instrument (HLS19-VAC), both developed by the Measuring Population and Organizational Health Literacy (M-POHL) Consortium. Data were analysed using IBM SPSS Statistics (v.29) with descriptive statistics, chi-square tests, Spearman’s correlation, and multiple linear regression. Mean HL score was 86.29 ± 22.04 and mean VL score was 18.29 ± 2.72. HL was significantly associated with gender (p < 0.001) and was the strongest independent predictor of physical activity (β = 0.735, R^2^ = 0.627, p < 0.001). VL was strongly associated with vaccination attitudes (r = 0.735, p < 0.001) but not with physical activity or smoking. HL and VL function as enabling rather than deterministic factors for health behaviour. Multi-component public health interventions combining HL promotion with environmental and policy-level strategies are needed.

## Introduction

Health literacy (HL) is widely recognized as a key determinant of health behaviors and outcomes. Based on previous research, HL can be consistently linked to preventive behaviors [1], health care utilization, and health decision making, and is defined as knowledge, motivation, and competences to access, understand, evaluate, and use health information in the context of health care, disease prevention, and health promotion.

Vaccine literacy (VL) is a more specific construct, defined as the ability to find, understand, evaluate and apply vaccine information to make informed vaccine decisions [2]. The evidence is growing that VL is a key determinant of attitudes and uptake of vaccinations [3, 4]. However, there have been limited studies investigating the co-occurrence of both HL and VL with general lifestyle behaviours such as physical activity and smoking.

In developed countries, higher HL is associated with an increased likelihood of physical activity and decreased likelihood of smoking [5]. However, there is conflicting evidence when socio-demographic factors are taken into account and weak evidence in low- and middle-income countries (LMICs). The challenges faced by Pakistan’s digital access, health system capacity, and sociocultural context can have a unique impact on these relationships [6, 7].

The present study aims to explore the prevalence of HL and VL among young adults in Pakistan, their association with socio-demographic factors, and their association with lifestyle factors (physical activity, smoking, and attitude towards vaccination). The study also examines whether HL and VL are independent predictors of physical activity after controlling for socio-demographic variables.

### Hypotheses

H_0_: There is no significant association between HL, VL, and lifestyle behaviours. H_1_: HL and VL are significantly associated with lifestyle behaviours.

## Materials and methods

### Study design and setting

An analytical, cross-sectional, quantitative study was carried out in Pakistan among adults aged 18 years and above. Data were gathered through a structured questionnaire administered either online (Google Forms) or paper-based. Data collection was conducted from August to October 2025. Ethical approval was granted by the Institutional Ethics Committee of Lithuanian Sports University (Approval No. SMTEK-75). All participants provided signed informed consent before participation, and confidentiality and anonymity were maintained throughout.

### Participants and sampling

Participants were recruited through non-probability convenience sampling (n = 587). The sample comprised primarily students and young adults. Inclusion criteria were: age ≥18 years, ability to understand and complete the questionnaire, and provision of informed consent. Participants with incomplete data were excluded from the final analysis. Results should be interpreted with respect to the sampled population rather than the general adult population of Pakistan.

### Instruments

Socio-demographic characteristics collected included age, gender, marital status, educational level, height, and weight.

#### Health literacy

HL was evaluated using the HLS19-Q instrument developed in the European Health Literacy Population Survey 2019–2021 (HLS19) by the M-POHL WHO Action Network [2]. The instrument measures how people can access, understand, appraise, and use health information in health care, disease prevention, and health promotion contexts [1]. The scale consisted of 17 items with a 5-point Likert scale (very difficult = 1 to very easy = 5). Raw item scores were transformed to a standardised 0–100 index following the HLS19 scoring formula, with higher scores indicating higher HL.

#### Vaccine literacy

VL was assessed using the HLS19-VAC instrument, also developed within the HLS19 project by the M-POHL Consortium [2]. The HLS19-VAC evaluates the knowledge, motivation, and skills needed to find, understand, evaluate, and apply vaccination-related information. It consisted of eight items rated on a five-point Likert scale summed to a composite VL score.

#### Physical activity

Physical activity was assessed by self-reported frequency of engagement in at least 30 minutes of moderate-to-vigorous activity per day. Responses were categorised as low (0–1 days/week), moderate (2–4 days/week), or high (≥5 days/week).

#### Smoking behaviour

Smoking was assessed through self-reported items on frequency and quantity of tobacco use, categorised as low (non-smokers or occasional smokers), moderate (<10 cigarettes/day), or high (≥10 cigarettes/day).

#### Vaccination attitudes

Five structured questions assessed attitudes regarding vaccine importance, safety, efficacy, disease prevention, and religious alignment. Responses were recorded on a 4-point Likert scale (strongly agree to strongly disagree) and a composite attitude score was computed.

### Statistical analysis

IBM SPSS Statistics (Version 29) was used for data analysis. Participant characteristics were summarised using descriptive statistics. Chi-square tests assessed associations between categorical variables; one-way ANOVA examined HL differences across age groups; independent samples t-tests compared mean scores by gender. Spearman’s correlation assessed relationships between HL, VL, and lifestyle variables. Multiple linear regression examined predictors of physical activity using HL, VL, gender, age, and education as predictor variables. A p-value of <0.05 (two-tailed) was considered statistically significant. The study was reported in accordance with the Strengthening the Reporting of Observational Studies in Epidemiology (STROBE) guidelines [8].

## Results

### Socio-demographic characteristics

587 participants were analysed (52.0% male, 48.0% female). Mean age was 21.54 ± 6.76 years. The majority were single (86.7%) and had high school education (64.7%). Mean HL score was 86.29 ± 22.04 on the standardised 0–100 scale and mean VL score was 18.29 ± 2.72 (Table 1).

**Table 1.** Socio-demographic characteristics and descriptive statistics of study participants.

| Characteristic | Category | n (%) | Mean ± SD |
| --- | --- | --- | --- |
| Gender | Male | 305 (52.0%) | — |
|  | Female | 282 (48.0%) | — |
| Age (years) | — | — | $21.54 \pm 6.76$ |
| Marital status | Single | 509 (86.7%) | — |
|  | Married | 75 (12.8%) | — |
| Education | High school | 380 (64.7%) | — |
|  | Bachelor's | 152 (25.9%) | — |
|  | Master's | 44 (7.5%) | — |
|  | Doctoral | 11 (1.9%) | — |
| HL score (0–100) | — | — | $86.29 \pm 22.04$ |
| VL score (raw) | — | — | $18.29 \pm 2.72$ |

### Health literacy scores by gender and age group

An independent samples t-test confirmed significantly higher mean HL scores in males (mean difference = 7.67, 95% CI: 4.12–11.23; t(545.43) = 4.243, p < 0.001; Levene’s F = 15.631, p < 0.001, equal variances not assumed). No significant difference in HL scores was found between age groups (F(2, 489) = 1.427, p = 0.241).

### Physical activity and smoking by gender

A significant difference in physical activity level was found between males and females (χ^2^(2) = 48.38, p < 0.001). Males had higher rates of high physical activity (39.7%) compared with females (14.2%). There was no significant gender difference in smoking level (χ^2^(2) = 5.53, p = 0.063) (Tables 2 and 3).

**Table 2.** Physical activity levels by gender.

| Physical activity level | Male n (%) | Female n (%) | Total n (%) |
| --- | --- | --- | --- |
| Low | 13 (4.3%) | 22 (7.8%) | 35 (6.0%) |
| Moderate | 171 (56.1%) | 220 (78.0%) | 391 (66.6%) |
| High | 121 (39.7%) | 40 (14.2%) | 161 (27.4%) |
| Total | 305 (52.0%) | 282 (48.0%) | 587 (100%) |

<sup>a</sup>Note: $\chi^2(2) = 48.38, p < 0.001$ .

**Table 3.** Smoking behaviour levels by gender.

| Smoking level | Male n (%) | Female n (%) | Total n (%) |
| --- | --- | --- | --- |
| Low | 78 (25.6%) | 97 (34.4%) | 175 (29.8%) |
| Moderate | 202 (66.2%) | 163 (57.8%) | 365 (62.2%) |
| High | 25 (8.2%) | 22 (7.8%) | 47 (8.0%) |
| Total | 305 (52.0%) | 282 (48.0%) | 587 (100%) |

Note: $\chi^2(2) = 5.53, p = 0.063$ .

### Vaccination attitudes

The majority of participants held positive vaccination attitudes. More than 90% agreed or strongly agreed that vaccines are important (95.6%), safe (94.0%), effective (89.7%), and prevent disease spread (93.0%). Agreement on religious compatibility was lower (81.1%), with 18.9% disagreeing or strongly disagreeing (Table 4).

**Table 4.** Distribution of attitudes toward vaccination.

| Statement | Agree/Strongly agree n (%) | Disagree/Strongly disagree n (%) |
| --- | --- | --- |
| Vaccines are important to protect myself and my children | 561 (95.6%) | 26 (4.4%) |
| Vaccines are safe | 552 (94.0%) | 35 (6.0%) |
| Vaccines are effective | 526 (89.7%) | 61 (10.4%) |
| Vaccination prevents disease spread | 546 (93.0%) | 41 (7.0%) |
| Vaccination fits religious beliefs | 476 (81.1%) | 111 (18.9%) |

### Correlation analysis

HL showed a strong positive correlation with physical activity (r = 0.717, p < 0.001) and a weak negative correlation with smoking (r = −0.111, p = 0.007). VL was strongly associated with vaccination attitudes (r = 0.735, p < 0.001) but did not significantly correlate with physical activity or smoking (Table 5).

**Table 5.**
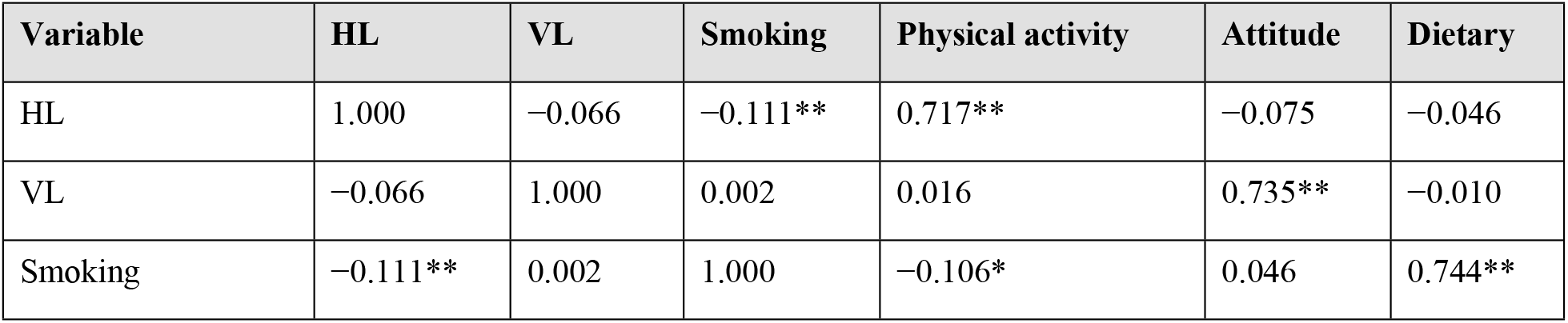
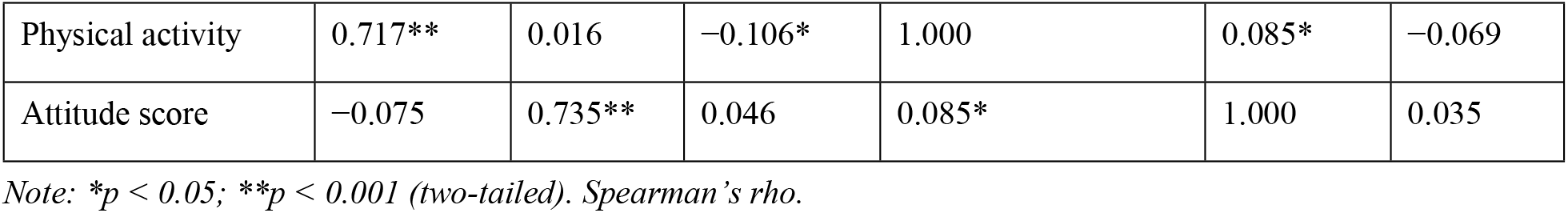
Spearman’s correlation matrix: HL, VL, and lifestyle variables (n = 587).

| Variable | HL | VL | Smoking | Physical activity | Attitude | Dietary |
| --- | --- | --- | --- | --- | --- | --- |
| HL | 1.000 | -0.066 | -0.111** | 0.717** | -0.075 | -0.046 |
| VL | -0.066 | 1.000 | 0.002 | 0.016 | 0.735** | -0.010 |
| Smoking | -0.111** | 0.002 | 1.000 | -0.106* | 0.046 | 0.744** |
| Physical activity | 0.717** | 0.016 | −0.106* | 1.000 | 0.085* | −0.069 |
| Attitude score | −0.075 | 0.735** | 0.046 | 0.085* | 1.000 | 0.035 |

Note: \* $p < 0.05$ ; \*\* $p < 0.001$ (two-tailed). Spearman's $\rho$ .

### Multiple linear regression: predictors of physical activity

A statistically significant multiple linear regression model was identified (F(5,581) = 195.584, p < 0.001, R^2^ = 0.627), explaining 62.7% of the variance in physical activity. HL was the strongest independent predictor (β = 0.735, p < 0.001). Gender, age, and VL were also significant predictors; education was not statistically significant (Table 6).

**Table 6.** Multiple linear regression: predictors of physical activity.

| Predictor | B | SE | $\beta$ | p-value | 95% CI |
| --- | --- | --- | --- | --- | --- |
| Constant | 36.258 | — | — | < 0.001 | — |
| Health literacy | 0.147 | — | 0.735 | < 0.001 | — |
| Vaccine literacy | 0.126 | — | 0.078 | 0.003 | — |
| Gender | −1.520 | — | −0.172 | < 0.001 | — |
| Age | −0.063 | — | −0.097 | 0.002 | — |
| Education | −0.151 | — | −0.024 | 0.416 | — |

Note: Dependent variable = self-reported days per week with $\geq 30$ min physical activity. $R^2 = 0.627$ , Adjusted $R^2 = 0.624$ , $F(5, 581) = 195.584$ , $p < 0.001$ .

## Discussion

This cross-sectional study explored HL and VL among young adults in Pakistan using the HLS19 instruments developed by the M-POHL Consortium. The mean HL score (86.29 ± 22.04) indicated overall high HL in this sample. The mean VL score was 18.29 ± 2.72.

### Levels of health and vaccine literacy

The high mean HL score may reflect the predominantly young, educated sample. Compared with Lithuanian adolescents evaluated using the same instruments who showed notably higher scores [4], the Pakistani sample reflects the influence of different sociocultural and healthcare environments.

### Gender differences in health literacy

A significant gender difference was found, with males scoring significantly higher than females (mean difference = 7.67, 95% CI: 4.12–11.23; p < 0.001), consistent with previous studies in Pakistan and other developing countries [7]. Caution is required given the convenience sampling design; gender differences may reflect sociocultural norms in health information seeking rather than capability differences.

### Health literacy as an independent predictor of physical activity

HL was the strongest independent predictor of physical activity (β = 0.735, p < 0.001, R^2^ = 0.627), consistent with the strong bivariate correlation (r = 0.717). This association persisted after controlling for gender, age, and education, suggesting that HL exerts an independent effect beyond socio-demographic factors. These findings align with the broader literature linking HL to health-promoting behaviours [5, 9].

### Vaccine literacy and vaccination attitudes

VL was highly correlated with positive vaccination attitudes (r = 0.735, p < 0.001), consistent with European studies using the same HLS19-VAC instrument [3, 4]. No association was found between VL and physical activity or smoking, confirming that VL is a domain-specific construct. A lower level of agreement on religious compatibility (81.1%) underscores the importance of culturally competent health communication in the Pakistani context, where religious factors play a significant role in health decision-making.

### Limitations

Several limitations should be noted. First, the cross-sectional design does not allow causal conclusions. Second, convenience sampling limits generalisability. Third, self-reported data may introduce recall bias and social desirability bias. Fourth, the HLS19-VAC and HLS19-Q instruments were originally designed and validated for European populations; cross-cultural applicability in Pakistan has not been formally assessed. Fifth, some chi-square analyses had low expected cell counts and should be interpreted with caution.

## Conclusions

This cross-sectional study assessed HL and VL using HLS19 instruments among young adults in Pakistan. HL was strongly associated with physical activity and remained a significant independent predictor after adjustment for socio-demographic covariates. VL was strongly linked to vaccination attitudes but not to general lifestyle behaviours. HL and VL function as enabling rather than deterministic factors for health behaviour, shaped by a complex interplay of cognitive, social, environmental, and structural influences. Public health interventions should adopt multi-component approaches combining HL promotion with targeted environmental and policy-level strategies. Future research should employ longitudinal designs and formally validated locally adapted instruments.

## Data Availability

The data underlying this study cannot be made publicly available due to ethical restrictions and participant confidentiality commitments made at the time of data collection. Data may be made available to qualified researchers upon reasonable request to the corresponding author.

## Acknowledgments

The authors thank Lithuanian Sports University for institutional support in conducting this study.

## Supporting information

**S1 Checklist**. STROBE checklist for observational studies.

## Author contributions

Conceptualisation, S.M.; methodology, S.M. and K.M.; formal analysis, S.M.; validation, K.M.; writing—original draft preparation, S.M.; writing—review and editing, S.M. and K.M.; project administration, S.M. All authors have read and agreed to the published version of the manuscript.

## Data availability statement

The data that support the findings of this study are available from the corresponding author upon reasonable request. Data will be deposited in a public repository (Dryad or Zenodo) upon acceptance.

## Ethics statement

This study was carried out in accordance with the recommendations of the Institutional Ethics Committee of Lithuanian Sports University (Approval No. SMTEK-75) and with the principles expressed in the Declaration of Helsinki. All participants provided written informed consent prior to participation.

## AI use statement

The authors acknowledge the use of ChatGPT (OpenAI) for grammar checking, language refinement, and sentence structure improvement during manuscript preparation. The authors have reviewed and revised all suggestions and are fully responsible for the accuracy and integrity of the work.

